# Effects of Physical Exercises on Emotion Regulation: A Meta-Analysis

**DOI:** 10.1101/2022.07.04.22277120

**Authors:** Jie Liu, Shuqing Gao, Liancheng Zhang

## Abstract

**Background:** At present, there are inconsistent results in the research on the effect of physical exercise on emotion regulation ability, and there is no relevant research on how to exercise scientificlc. Therefore, on the basis of previous research, this study conducted a meta-analysis on the theme of physical exercise affecting emotion regulation ability according to the statement of PRISMA, and added relevant moderator variables to clarify the effect of physical exercise on emotion regulation ability.

**Objective:** We identified physical exercise studies and randomized controlled trials (RCTs) of emotion regulation ability that reported overall effect, heterogeneity, and publication bias of physical exercises on emotion regulation ability.

**Methods:** We searched for RCT s of exercise interventions on emotion regulation ability from databases including PubMed, Web of Science, Ebsco, The Cochrane Library, CNKI (China National Knowledge Infrastructure), and Wanfang, from January 1 2000 to January 1 2022. We performed methodological quality evaluations on the included literature and graded evidence with a meta-analysis using Review Manager 5.3 software.

**Results:** In total, 10 RCT s were included; the overall results of the meta-analysis (936participants) indicated that physical exercises improved emotion regulation ability (standardized mean difference (SMD) = 0.47; 95% confidence interval (CI) 0.21 to 0.72; p < 0.05), sensory arousal ability (SMD = 0.70; 95% CI 0.14 to 1.27; p < 0.05), and emotion regulation strategies (SMD = 0.46; 95% CI 0.04 to 0.87; p < 0.05); Subgroup analysis showed that a single exercise of more than 30 minutes can improve emotional regulation ability, which is not affected by exercise intensity, period and the health status of the participants.

**Conclusion:** Physical exercises can effectively improve the emotion regulation ability. The effects are considered as moderate effect sizes. While single exercise time should be at least 30 minutes.Limited by the quantity and quality of the included studies, the above conclusions need to be verified with more high-quality studies.

PROSPERO registration number: CRD42021217978

## INTRODUCTION

Emotion regulation is the process of the occurrence, experience and expression of individual emotional changes, which can be carried out not only at the explicit level, but also as an implicit process (Gross, 2013). According to Gross’s process model, emotion regulation includes four stages: (1) feeling emotion, (2) attention, (3) evaluation, (4) reaction (Gross, 1998), that is, each stage of emotion regulation model is affected by multiple cognitive control processes, which suggests that there is a close relationship between emotion and emotion regulation, but they are not equivalent. Emotion regulation is a dynamic process, and disorders may occur at any stage of the process. Therefore, the level of emotion regulation ability plays an important role in improving our emotions. People who are ineffective at regulating their emotions experience many distressing periods, which may evolve into diagnosable depression or anxiety (Mennin et al., 2007; Nolen-Hoeksema, Wisco, and Lyubomirsky, 2008). This shows that successful emotion regulation is a prerequisite for adaptive function (Gross, 1999), and improving emotion regulation ability is an important way to prevent related emotional diseases and promote healthy life.

At a time when the concept of “exercise is good medicine” prevails, there are a lot of studies on the effect of physical exercise on improving emotions. So can physical exercise also improve the ability to regulate emotions? Some studies have suggested that physical exercise has a positive impact on emotion regulation ability

(Brown et al., 2013; Giles et al., 2017), of which aerobic exercise intervention has a more significant effect (Bernstein and McNally, 2017; Edwards, Rhodes, and Loprinzi, 2017). Similarly, Augustine and Hemenover (2009) also found that aerobic exercise is one of the effective emotion regulation intervention strategies, with an improved effect size of 0.47. Some scholars have proposed that moderate aerobic exercise lasting 20-30 minutes may be the most effective for improving emotional regulation (Berger and Motl, 2000). However, some studies have also found that physical exercise has no significant or negative impact on emotional regulation. For example, some scholars have explored the improvement of emotional regulation by a single session of aerobic exercise and meditation, and the results have shown no significant difference (Edwards et al., 2018), and some scholars have proposed that when the load of exercise exceeds the ventilation threshold, the pleasure will decrease, which will negatively affect the ability of emotion regulation (Ekkekakis, Parfitt, & Petruzzello, 2011).

To sum up, there are still inconsistent results in the research on the effect of physical exercise on emotion regulation ability, and there is no relevant research on how scientific exercise can improve emotion regulation ability. Therefore, on the basis of previous research, this study conducted a meta-analysis on the theme of physical exercise affecting emotional regulation ability according to the statement of PRISMA, and added relevant moderator variables to clarify the impact of physical exercise on emotional regulation ability, and provide exercise guidance to promote public emotional well-being.

## METHOD

Our research follows the requirements of the international meta-analysis writing guidelines (the PRISMA statement for reporting systematic reviews and meta-analyses of studies that evaluate health-care interventions: explanation and elaboration) for selecting and utilizing research methods (Shamseer et al., 2015).

### Retrieval Strategy

The databases PubMed, Web of Science, Ebsco, The Cochrane Library, China National Knowledge Infrastructure (CNKI), and Wan Fang were searched from January 1 2000 to January 1 2022. Two reviewers independently searched articles published in Chinese and English, supplemented by a manual search, and retrospectively included references if necessary. The following two sets of search terms were used: “physical activity” or “exercises” or “physical fitness” or “physical endurance” or “motor activity” or “physical education” or “sport” or “basketball” or “football” or “running” or “cycling” or “jumping” or “dancing” or “tai chi” or “yoga” or “aerobic” and “emotion regulation” or “emotion regulation strategies” or “emotion regulation difficulties” or “emotion regulation ability” or “emotion management.” If an article was incomplete or unavailable, we contacted the corresponding author by email to obtain detailed information. For literature tracing, based on the retrieved literatures or related references listed in the review, we used Baidu Scholar and Google Scholar to search for them retrospectively.

### Inclusion Criteria and Exclusion Criteria

Two reviewers independently screened the articles. When there was a disagreement between the two reviewers, a third reviewer evaluated the original study to reach a consensus. Any potentially relevant research needed to meet the following inclusion criteria: (1) the exercise group was the primary intervention measure (e.g., aerobic-based, motor skill-based, combining aerobic, muscular activity, yoga, basketball), compared with different types of control groups (i.e., no-intervention control group, waiting list, and routine care) and all the intervention measures were motor skill-based or aerobic-based and clearly defined in terms of the exercise protocol; (2) preliminary studies were randomized controlled trials, and the randomization was done either on an individual or on a group (e.g., classroom) basis; (3) outcome indicators included test data on emotion regulation, with a minimum of one outcome with quantitative data for calculating the pooled effect size. Conditions for exclusion from the study included: (1) ambiguous explanations of exercises interventions; (2) irrelevant outcomes; and (3) studies for which the full text could not be obtained.

### Data Extraction

Two reviewers independently extracted data according to a predefined protocol. If there were any differences or inconsistencies, they would discuss the study with a third reviewer. They gathered the following information: (1) literature information, including author name, year of publication; (2) sample size; (3) socioeconomic status; (4) basic information of subjects, mainly used to divide the type of population; (6) intervention program; (7) measurements; (8) adverse events and follow-up.

### The Methodological Quality of the Included Studies

The literature quality of the included literature was assessed by Review Manager 5.3. The 11 literatures studied were comprehensively scored in six aspects: (1) sequence generation (whether randomization was used); (2) whether allocation concealment was performed and whether the allocation method was correct; (3) whether blinding was implemented; (4) whether there was reporting errors. (5) whether there is a selective outcome report; (6) whether there is any other bias report. According to the recommendations of evidence-based medicine research guidelines, two panelists used the “Risk of bias assessment” tool of the Cochrane systematic review to assess included studies (Higgins et al., 2019), and any differences were resolved by a third reviewer.

### Statistical Analysis

Review Manager 5.3 was used as data processing software. We used the standardized mean difference (SMD) with a 95% confidence interval (CI) to analyze the combined effect size. According to the Cochrane systematic review manual, if a study included more than one control group, the sample size of the exercise intervention should be equally assigned during pair comparison in order to avoid analysis unit errors (Handoll, Howe, and Madhok, 2002). If statistical heterogeneity was found across studies (I^2^ ≥ 50%, *p* < 0.10), we applied the random-effects model—otherwise, the fixed-effect model was applied—and we used the Hedges’ g method to reflect the magnitude of exercise intervention (Liu et al., 2019). According to the criteria for evaluating effect volumes, a small effect was between ≥0.2 and <0.5, a medium effect was between ≥0.5 and <0.8, and a large effect was ≥0.8 (Hanley et al., 2003). The heterogeneity of the included studies was determined with the p-value (threshold point of 0.1) and I^2^ statistics (25, 50, and 75%, representing small, medium, and large heterogeneity, respectively) (Liu et al., 2019).

Given that overall effect sizes may be influenced by heterogeneity factors, subgroup analyses were performed for participant’s health, intervention duration, intensity, and time: (1) healthy people vs. unhealthy people; (2)medium intensity vs. low intensity; (3) acute exercise vs. long-term exercises (movements with relatively stable sports environments, such as yoga and running) (Liu et al., 2019); (4) intervention time referred to the timing of intervention, where a threshold of 30 min for exercise was recommended.

## RESULTS

### Literature Search Results

The latest review of electronic searches (as of January 2022) retrieved a total of 3914 articles. During the preliminary screening, we excluded 3229 studies based on their title and abstract for reasons including duplications (n = 649), language (n = 17), or not being related to the subject content (n = 2563). Further screening was performed by reading the full text, and 674 records were excluded because of non-randomized controlled trials (n = 512), no reported data for analysis (n = 57), review (n = 59), or no major exercise interventions (n = 46). Finally, the meta-analysis included 11 articles (Figure 1).

**FIGURE 1.**
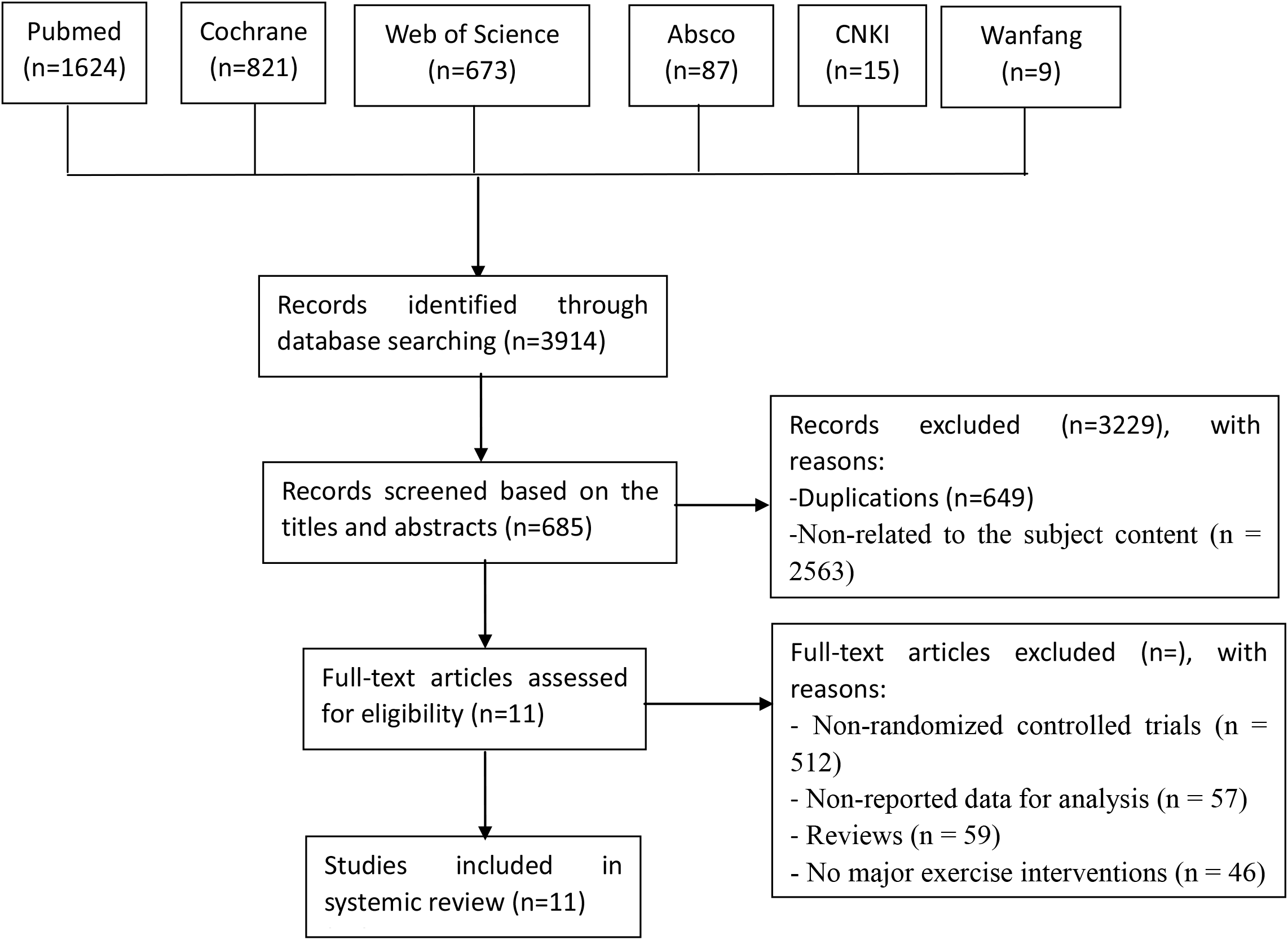
Flow of the study selection method

### Eligible Research Features

The 11 articles included were randomized controlled trialsc (Bernstein and McNally, 2017; Tse, 2020; Stolarska et al., 2019; Bernstein and McNally, 2018; Bernstein, Heeren, and McNally, 2020; Alderman et al., 2016; Bahmani et al., 2020; Giles et al., 2018; Edwards et al., 2018; Zhang et al., 2017; 2021), including 936 participants, of which 455 were in the experimental group and 447 were in the control group (Tables 1).

**TABLE 1.**
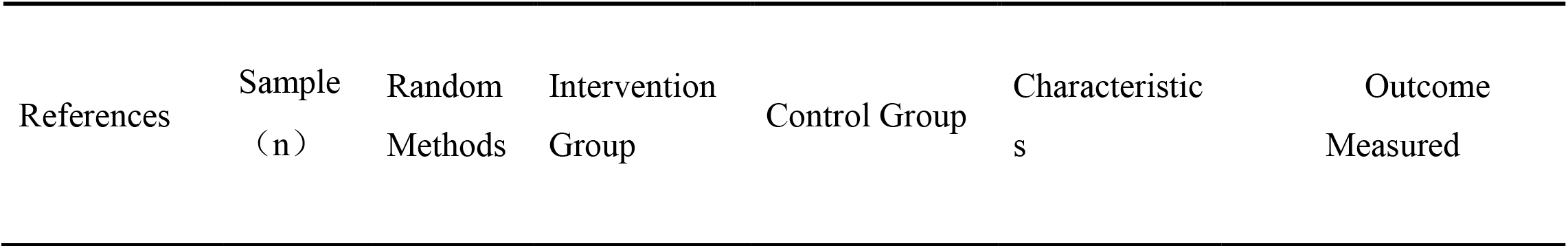
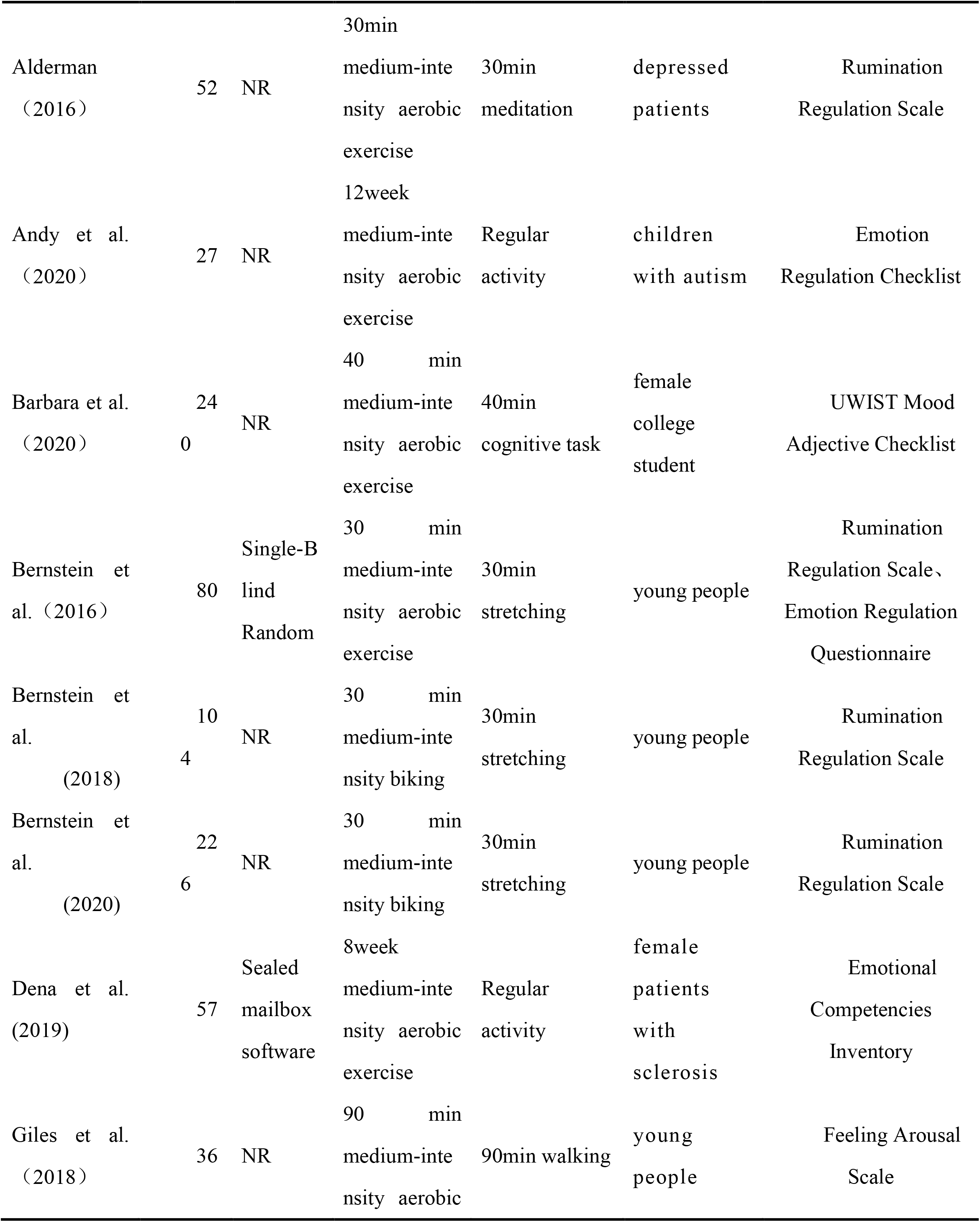
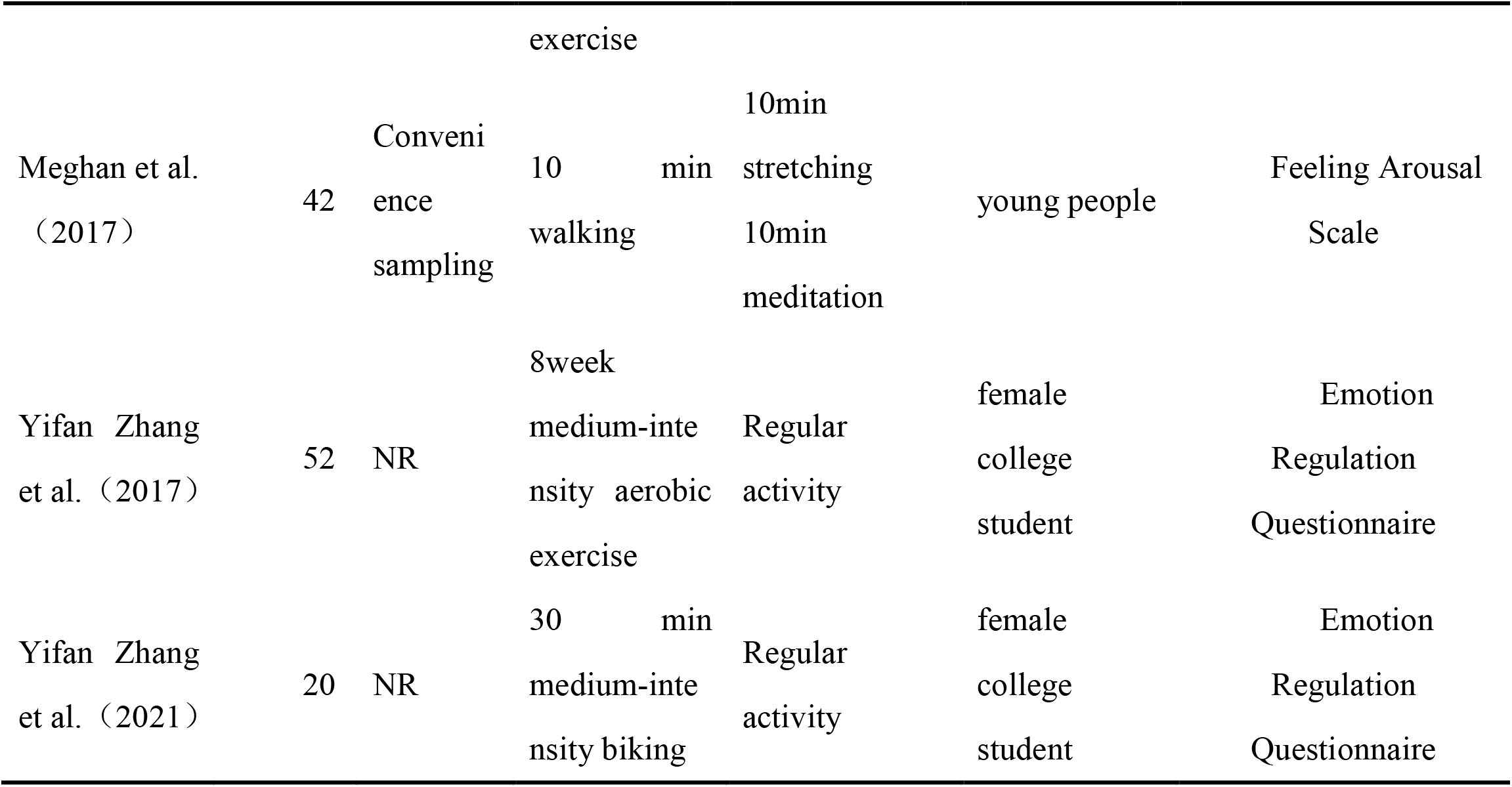
Characteristics of the studies included in the meta-analysis

### Methodological Quality Evaluation

The overall risk of bias of the included studies was low because exercise required exercise instruction for participants and could not be blinded, but this did not have a significant impact on outcome assessment. Specifically as shown in Figure 2.

**FIGURE 2.**
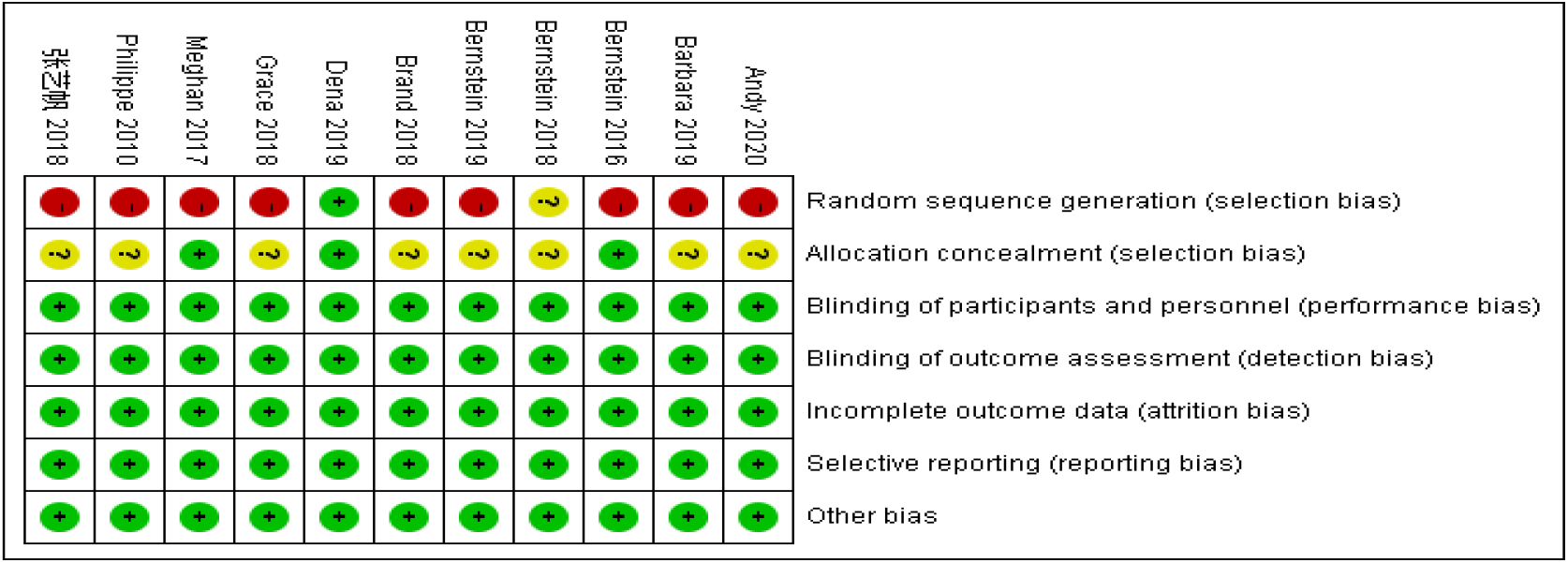
Risk of Bias Proportional Plot

### Publication Bias

The testing methods for publication bias mainly include: funnel plot, Begger’s test, and Egger’s test. Some scholars proposed that the funnel plot is a common method for measuring publication bias, and it is suitable for studies with continuous variables as outcome indicators, and the number of included studies is at least ten (Higgins, Green, and Scholten, 2008). To meet the requirements, this study used the funnel plot suggested by the Cochrane Library to conduct publication bias testing. The results showed that 11 experimental studies basically formed a funnel plot, as shown in Figure 3. Subsequently, the researchers used Egger’s test to conduct a secondary test, and the results showed that *p*=0.21<0.05, which indicates that the publication bias is not obvious and the data reliability is high.

**FIGURE 3.**
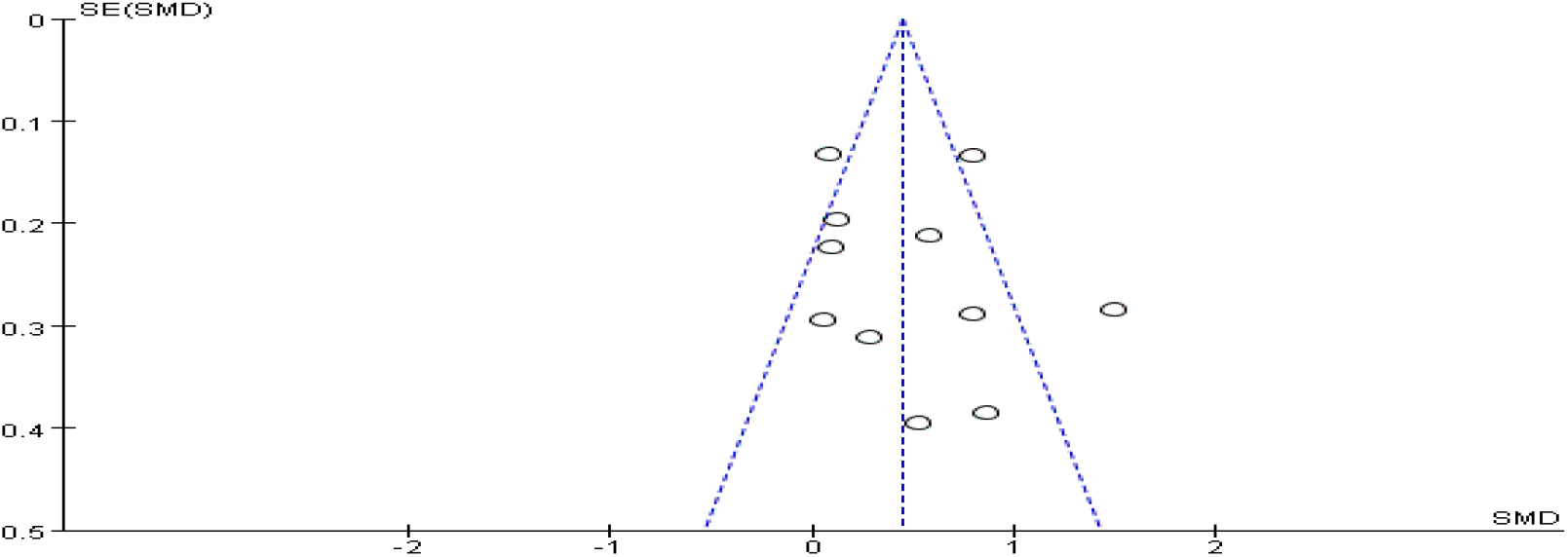
Funnel Plot of Publication Bias

## Meta-Analysis Results

### Physical Exercise and Emotional Regulation

11 studies examined the effect of exercises on emotion regulation. After analysis, this article has a high degree of heterogeneity (I^2^=91%), and then a sensitivity analysis was performed. The results showed that after excluding the study of Alderman et al., I^2^=67%, which is moderate heterogeneous. Due to the different scales, a random effect model was used to conduct a Meta-analysis of the final 10 references. The aggregated result showed a significant benefit in favor of exercises on emotion regulation ability (SMD = 0.47; 95% CI 0.21 to 0.72, *p* = 0.00) (Figure 4).

**FIGURE 4.**
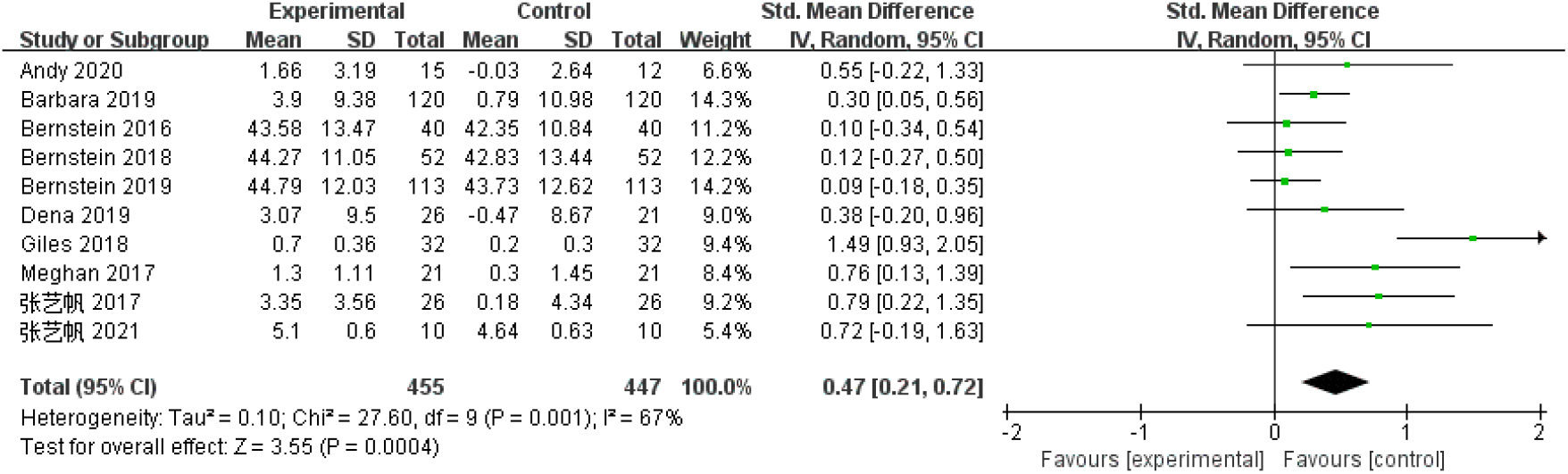
The Effect of Exercises on Emotion Regulation Ability

Effect sizes were quantified as large (SMD > 0.8), medium (0.5 < SMD < 0.8), or small (0.2 < SMD < 0.5) as suggested by Cohen (2013). This suggests that physical exercise can have a moderate effect on emotion regulation. In order to further explore the impact of physical exercise on different dimensions of emotion regulation ability, this paper divides emotion regulation ability into three aspects according to the Gross emotional process model: sensory arousal, rumination regulation, and emotion regulation strategies (including cognitive reappraisal and expression inhibition).

(1) Exercise and Rumination Regulation

4 studies (including 440 participants) explored the effect of physical exercise on rumination regulation. There was no heterogeneity among the studies (I^2^ = 0%), and the fixed effect model was used for analysis. The aggregated result showed there were no significant differences in rumination regulation (SMD =-0.13; 95% CI -0.31 to 0.06, *p* = 0.18) (Figure 5).

**FIGURE 5.**
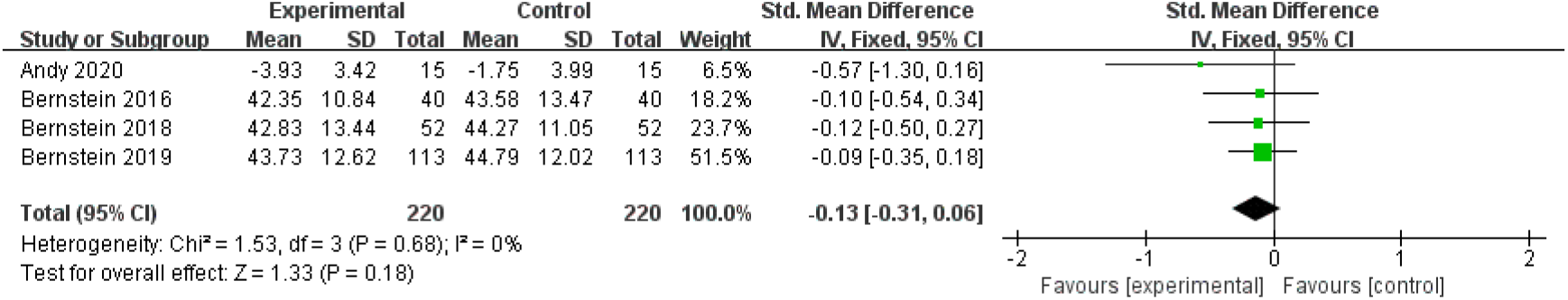
Evaluation of Rumination Regulation

(2) Exercise and Sensory Arousal

3 studies (including 346 participants) explored the effect of physical exercise on sensory arousal. There was moderate heterogeneity (I^2^ = 78%) among the studies. The random effects model was used for analysis. The aggregated result showed a significant benefit in favor of exercises on sensory arousal (SMD =0.70; 95% CI 0.14 to 1.27, *p* = 0.01) (Figure 6).

**FIGURE 6.**
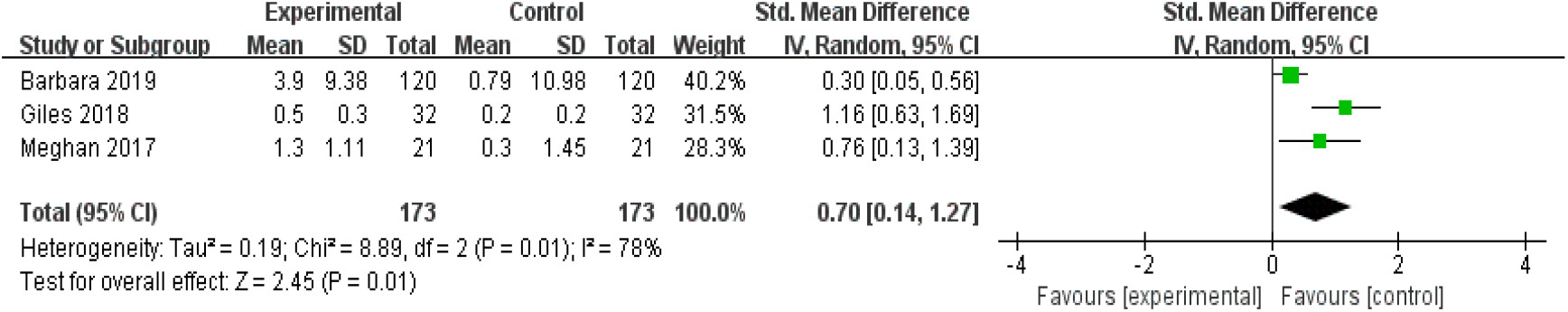
Evaluation of Sensory Arousal

(3) Exercise and Emotion Regulation Strategies

6 studies (including 305 participants) explored the effect of physical exercise on emotion regulation strategies. There was low heterogeneity (I^2^ = 42%) among the studies. The random effects model was used for analysis. The aggregated result showed a significant benefit in favor of exercises on emotion regulation strategies (SMD =0.46; 95% CI 0.04 to 0.87, *p* = 0.03) (Figure 7).

**FIGURE 7.**
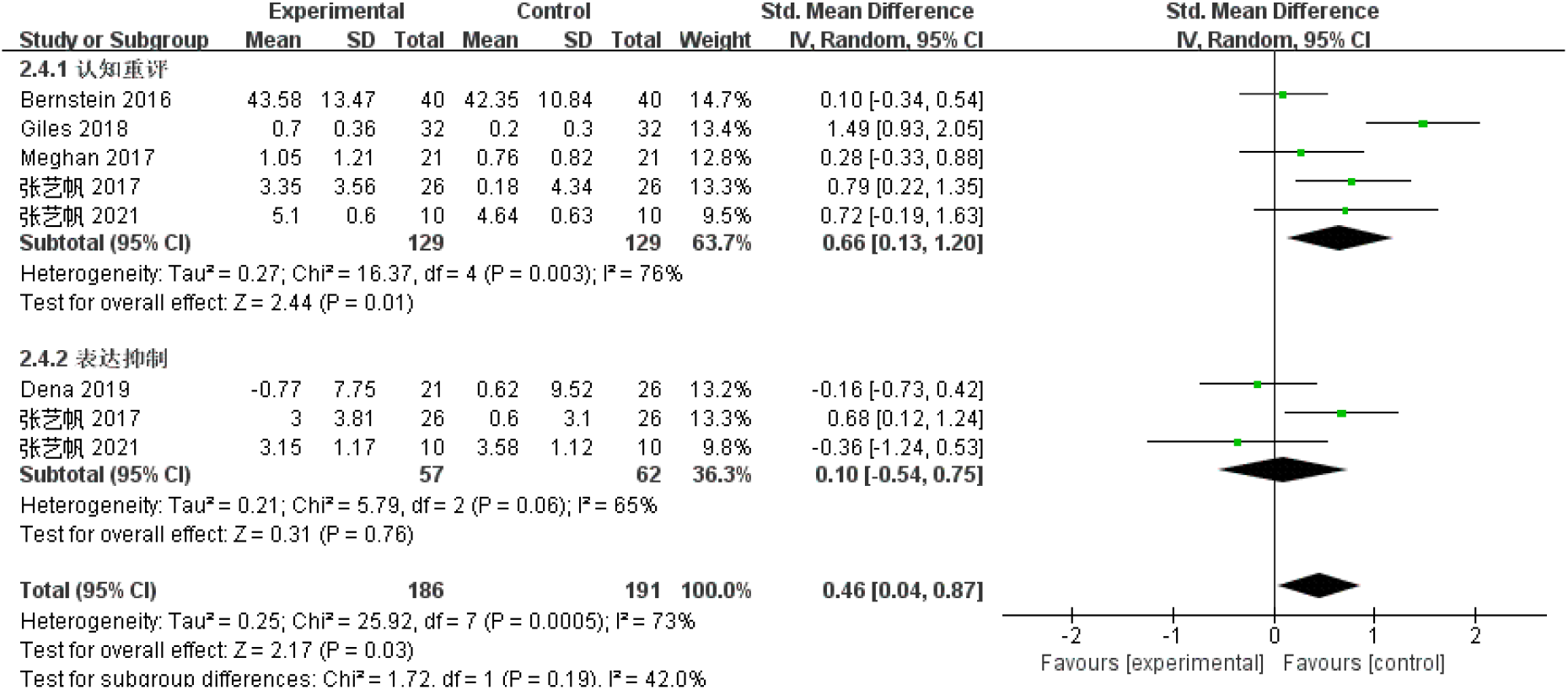
Evaluation of Emotion Regulation Strategies

### Subgroup Analysis

In addition to examining the effect of physical exercise on emotion regulation ability and its various aspects in general, this study also examined moderator variables that affect the relationship between the two. The four variables of intervention intensity, intervention duration, intervention period, and participants’ health were selected to carry out the moderating effect test.

(1) Exercise Intensity and Emotion Regulation

In terms of the exercise intensity, either moderate-intensity physical exercise interventions or low-intensity physical exercise interventions were employed in the original studies. 7 studies (including 749 participants) explored the effect of acute exercise on emotion regulation ability, and 3 studies (including 148 participants) explored the effect of long-term exercise on emotion regulation ability. There was moderate heterogeneity (I^2^ = 66%) among the studies. A random-effects model was used for analysis. The aggregated result showed there were no statistically significant differences on the ESs between the two types of interventions (*p* = 0.44) (Figure 8). It shows that regardless of the intervention of the exercise intensity, the emotion regulation ability of the participants who received physical exercise was higher than that of the participants who did not receive physical exercise.

**FIGURE 8.**
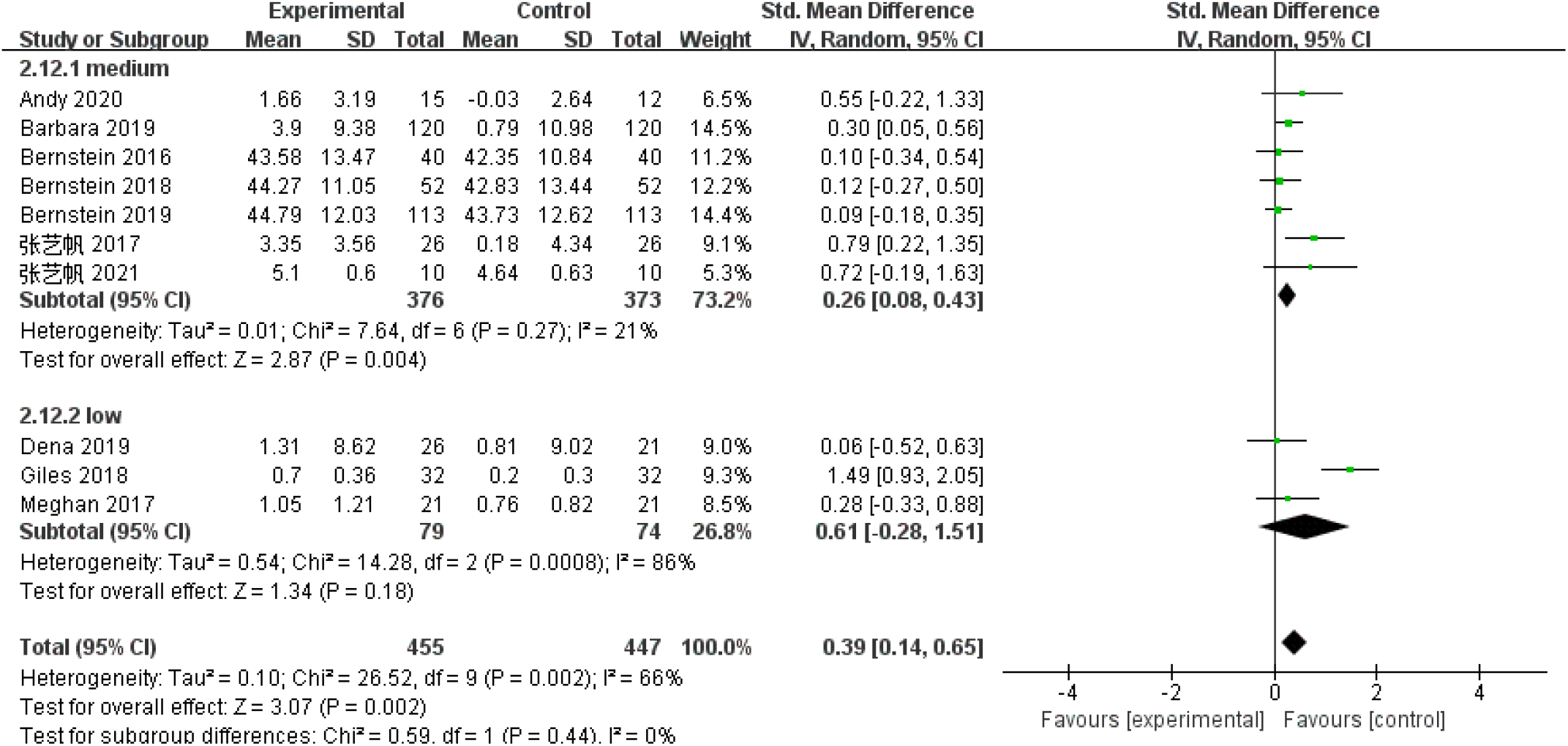
Evaluation of the Effect of Intervention Intensity on Emotion Regulation

(2) Exercise Period and Emotion Regulation

In terms of the exercise period, either acute exercise interventions or long-term exercise interventions were employed in the original studies. 7 studies (including 776 participants) explored the effect of moderate-intensity physical exercise on emotion regulation ability, and 3 studies (including 126 participants) explored the effect of moderate-intensity physical exercise on emotion regulation ability. There was moderate heterogeneity (I^2^ = 67%) among the studies. A random-effects model was used for analysis. The aggregated result showed there were no statistically significant differences on the ESs between the two types of interventions (p = 0.58) (Figure 9). It shows that regardless of the intervention of the exercise period, the emotion regulation ability of the participants who received physical exercise was higher than that of the participants who did not receive physical exercise.

**FIGURE 9.**
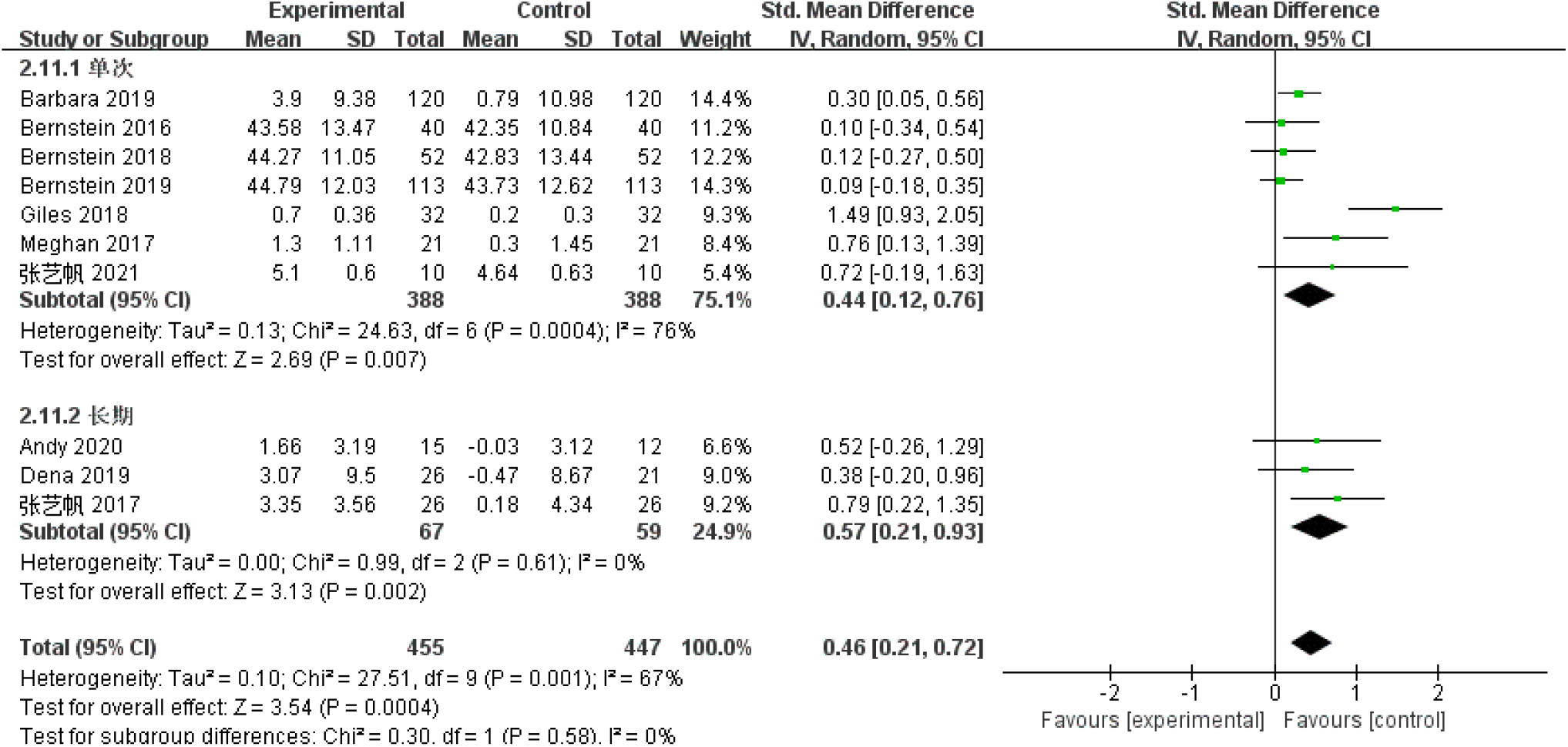
Evaluation of the Effect of Intervention Period on Emotion Regulation

(3) Participants and Emotion Regulation

In terms of the participants, either healthy people or unhealthy people were employed in the original studies. 7 studies (including 588 participants) explored the effect of physical exercise on the emotional regulation ability of healthy groups, and 2 studies (including 74 participants) explored the effect of physical exercise on the emotional regulation ability of unhealthy groups. There was moderate heterogeneity (I^2^ = 71%) among the studies. A random-effects model was used for analysis. The aggregated result showed there were no statistically significant differences on the ESs between the two types of interventions (p = 0.74) (Figure 10). It shows that regardless of the health status of the participants, the emotion regulation ability of the participants who exercised is higher than that of the participants who did not exercise.

**FIGURE 10.**
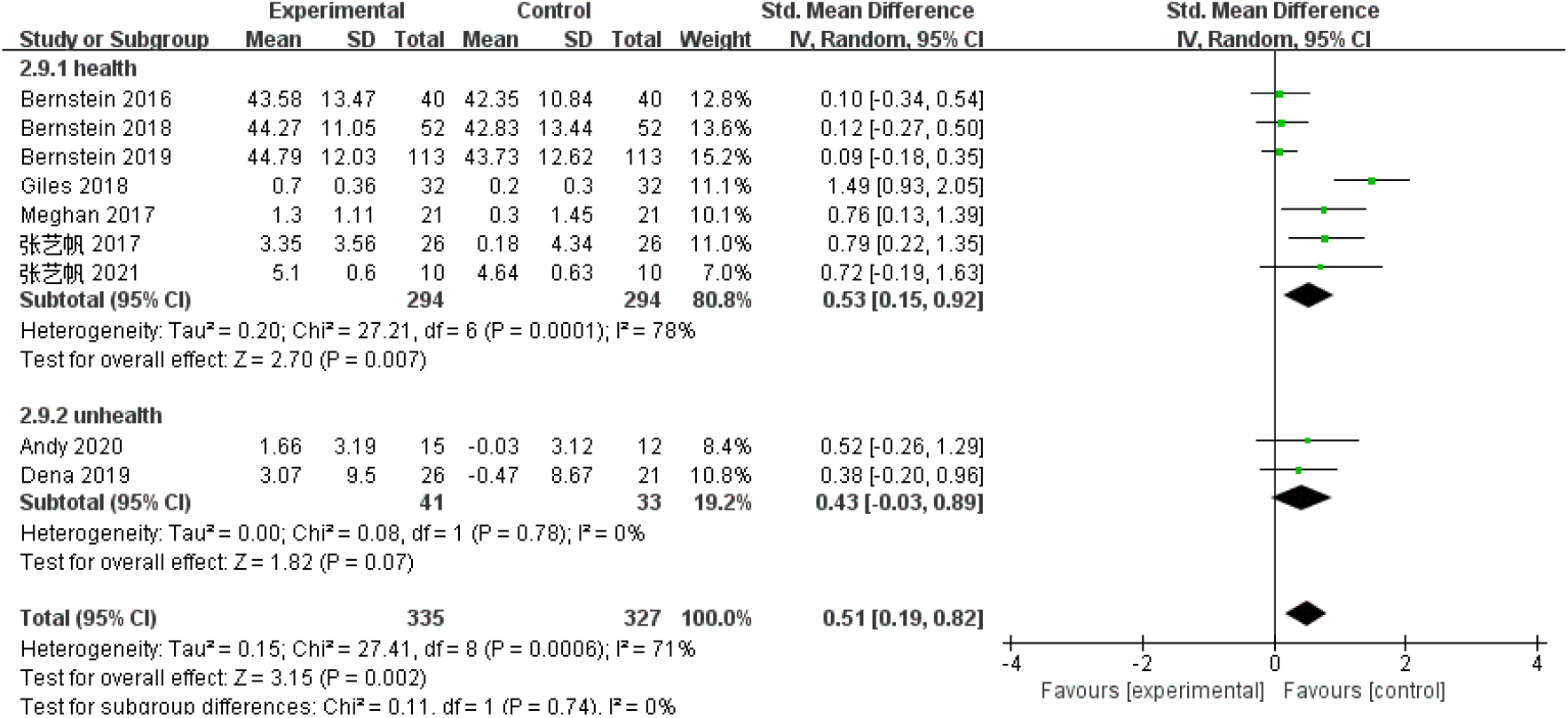
Evaluation of the Effect of Physical Exercise on Emotion Regulation Ability of Different Participants

(4) Exercise Duration and Emotion Regulation

In terms of the exercise duration, either session time (≤30 min) or prolonged exercise(>30 min) were employed in the original studies. 4 studies (including 452 participants) explore the effect of physical exercise under 30min on emotion regulation ability, and 2 studies (including 74 participants) explore the effect of physical exercise more than 30min on emotion regulation ability. There was moderate heterogeneity (I^2^ = 74%) among the studies. A random-effects model was used for analysis. The results showed that when the duration of a single intervention was less than 30min, physical exercise had no significant effect on emotion regulation ability (SMD =0.17; 95% CI -0.05 to 0.40, *p* = 0.13), while the duration of 30min and above has a significant effect on emotion regulation ability(SMD =0.80; 95% CI 0.21 to 1.39, *p* = 0.01), and the difference in effect value between the two groups is significant (*p* =0.00). This suggests that the duration of acute exercise should preferably be more than 30 min.

## Discussion

### The Validity of the Effect of Physical Exercise on Emotion Regulation Ability

Current studies on the effect of physical exercise on emotion regulation have reported inconsistent results. In order to explore the relationship between the two from an evidence-based perspective, this paper conducted a meta-analysis and integration of 10 studies that met the inclusion criteria. The model divides emotion regulation into three dimensions: rumination regulation, sensory arousal, and emotion regulation strategies. The results show that physical exercise can improve not only the emotion regulation ability but also sensory arousal and emotion regulation strategies significantly. So how does physical exercise affect emotion regulation?

This study integrates 10 included literatures to explore the path of physical exercise to improve emotion regulation ability. Among them, 7 literatures proposed that cognitive function is an effective mediating variable for exercise to improve emotion regulation ability. Activation of the cerebral cortex improves cognitive function and indirectly affects emotion regulation at different stages (Brown et al., 2013); 6 literatures suggested that physical exercise regulates hippocampal tissue by promoting the secretion of neurotransmitters (such as adrenal tissue and corticosterone in plasma). In turn, it directly affects the ability of emotion regulation; 4 literatures suggest that the sensory arousal enhanced by physical exercise will affect the physical and psychological state of individuals when they are awake, and directly improve the ability of emotion regulation; at the same time, the improvement of the level of arousal corresponds to the improvement of executive function significantly (Byun et al., 2014), and executive function plays a certain role in affecting emotion regulation ability (Garofalo, Battaglia, and di Pellegrino, 2019), which suggests that the action path of physical exercise affecting emotion regulation ability can be explained from three aspects, as shown in Figure 12.

**FIGURE 12.**
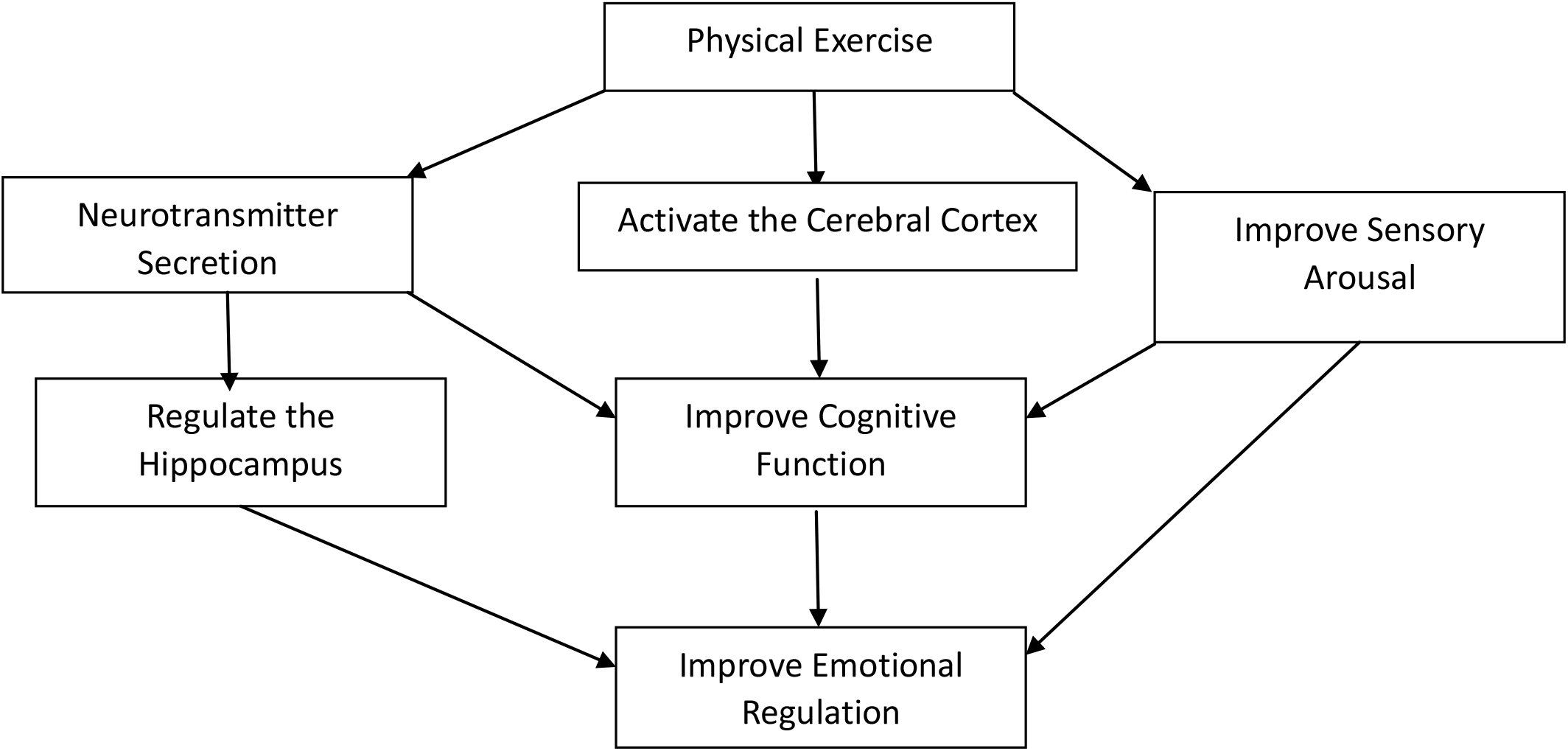
The Paths of Physical Exercise Affecting Emotion Regulation Ability

In conclusion, this paper provides a corresponding theoretical basis for physical exercise to improve emotion regulation ability, and provides a theoretical reference for the updating of emotion regulation models in the future.

### Moderator Variables of the Effect of Physical Exercise on Emotion Regulation Ability

To explore how to maximize the improvement of emotion regulation ability, this paper further included four moderator variables for subgroup analysis. The results showed that the intensity, period and participants of exercise did not affect the improvement of emotional regulation ability of physical exercise, which suggests that in the field of exercise behavior promotion, acute exercise can also have a positive impact on emotional regulation ability and improve physical and mental health. Individuals can choose exercise intensity according to their load tolerance and personal preference. In addition, the effect of the duration of acute exercise on the emotion regulation ability shows that more than 30 minutes’ exercise have a significant effect.

To sum up, whether it is a healthy group or a non-healthy group, physical exercise can have a significant positive effect on their emotional regulation ability. A single session of 30 or more minutes of low-to moderate-intensity exercise can improve emotional regulation, and it works even better in the long run. This conclusion can avoid the negative effects of excessive exercise or incorrect exercise, and then provide scientific exercise guidance for the greater improvement of emotion regulation ability.

### Limitations and Futures

Although the quality of the included literature is high, and the funnel plot of the included literature shows that there is no bias, the sample size of each study is not large enough, and the measurement methods of emotion regulation ability are different. These heterogeneity factors affected the results of the meta-analysis to a certain extent; secondly, due to the limited characteristics of the literature included in this paper, differences between aerobic and anaerobic, physical and mind-body, and individual and group exercise could not be compared, so there will be certain limitations in providing corresponding exercise programs.

Future research in this area could take a diverse approach to physical exercise, consider factors such as the exercise environment, and examine the dose-response nature of these effects. In addition, the data in the studies included in this paper are all recorded in the form of self-reports such as scales. In the future, the physiological indicators of emotion such as brain-derived neurotrophic factor heart rate and dopamine can be combined to comprehensively evaluate the changes in emotion regulation ability; or Combined with cognitive function, a model theory of emotion regulation process is proposed, and the internal mechanism of this positive effect is clarified.

## CONCLUSIONS

1. Physical exercise is an effective means to improve individual emotion regulation ability.
2. A single physical exercise of more than 30 minutes can improve the ability of emotional regulation, which is not affected by the exercise intensity, period and the health status of the intervention object, and the long-term adherence effect will be better.

## Data Availability

All data produced in the present study are available upon reasonable request to the authors. All data produced in the present work are contained in the manuscript

## DATA AVAILABILITY STATEMENT

The original contributions presented in the study are included in the article/supplementary material, further inquiries can be directed to the corresponding author.

## AUTHOR CONTRIBUTIONS

JL, SG, and LZ: conceptualization. JL and SG: methodology, software, and resources. SG and LZ: validation. JL, SG, and LZ : formal analysis. JL, SG, and LZ: investigation and data curation. JL, SG, and LZ: writing—original draft preparation. JL and SG : writing—review and editing. All authors contributed to the article and

approved the submitted version.

### FUNDING

This study was supported by China Social Science Foundation (No. 17BTY118) and Social Science Fund of the Ministry of Education of China (19YJCZH035)

## Notes

### Competing Interest Statement

The authors have declared no competing interest.

### Funding Statement

This study did not receive any funding

